# Assessing Facioscapulohumeral Muscular Dystrophy through Comparative Analysis of Bulk and Single-Cell Transcriptomes

**DOI:** 10.1101/2023.11.23.23298813

**Authors:** Saed Sayad, Mark Hiatt, Hazem Mustafa

## Abstract

**Background:** Facioscapulohumeral muscular dystrophy (FSHD) is a genetic disorder characterized by progressive weakening of the muscles. While the two types of FSHD (FSHD1 and FSHD2) have distinct genetic causes, they share similarities in their clinical presentations. Both result in muscle weakness, particularly in the face, shoulders, and upper arms. Genetic testing is essential for accurately diagnosing the specific type of FSHD and guiding treatment and management.

**Method:** We acquired bulk and single-cell gene expression data for FSHD2 from the *NIH portal website*. Our analysis involved an extensive array of differentially expressed genes, and pathway and gene ontology analysis. Using statistical tests, we identified the top up- and down-regulated genes, and the pathways and gene ontology terms characterizing those genes that exhibited substantial changes across both bulk and single-cell transcriptomes.

**Results:** The top 10 up-regulated genes identified in the bulk gene expression analysis represent a diverse range of biological functions, but all are associated with FSHD. In contrast to the bulk down-regulated genes, the single-cell top 10 down-regulated genes are primarily linked to muscle-related functions. These genes, such as *ACTC1, ACTA1, MYL11, MYH3, MYL6B, MYBPH, TPM2, MYL2, MYL1*, and *TNNI1* are integral to muscle contraction and skeletal muscle function. Moreover, all the top 10 single-cell down-regulated pathways are implicated in the pathogenesis of muscle dystrophy. Finally, the top 10 down-regulated gene ontology terms are all relevant to the pathogenesis of muscular dystrophy.

**Conclusions:** This study unequivocally demonstrates that single-cell transcriptomics surpasses bulk transcriptomics in elucidating the genes, pathways, biological processes, molecular functions, and cellular components associated with FSHD2. While bulk transcriptomics offers a broader perspective on gene expression, single-cell transcriptomics shines in its capacity to unveil cell-specific gene regulation, especially in the realm of muscle-related functions.

## Introduction

Facioscapulohumeral muscular dystrophy (FSHD) is a genetic disorder [1] characterized by progressive weakening of the muscles, primarily affecting the face, shoulder blades (scapulae), and upper arms (humeri). The two types of FSHD, types 1 and 2, each have different genetic causes. FSHD type 1 is associated with a deletion in a region on chromosome 4 called D4Z4; whereas FSHD type 2 is caused by mutations in a gene called *SMCHD1* which is a chromatin modifier necessary for the establishment and maintenance of CpG methylation of the inactive X chromosome and specific classes of repeated elements. In both FSHD1 and FSHD2, DNA unwinds and activates a dormant gene called *DUX4. DUX4*’s normal function is regulating an early stage of embryonic development, but its activation in adult muscle cells causes them to die prematurely. The precise underlying pathology of this condition remains yet to be fully understood. Bulk and single-cell transcriptomics are powerful techniques in understanding the molecular mechanisms and gene expression patterns associated with diseases. In this study, our primary emphasis is on discerning differentially expressed genes, as well as conducting pathway and gene ontology analysis in the context of FSHD2.

## Data

We acquired bulk (*GSE143453*) and single-cell (*GSE143452*) transcriptomes for FSHD2 from the NIH portal website (**Figures 1** and **2**).

**Figure 1:**
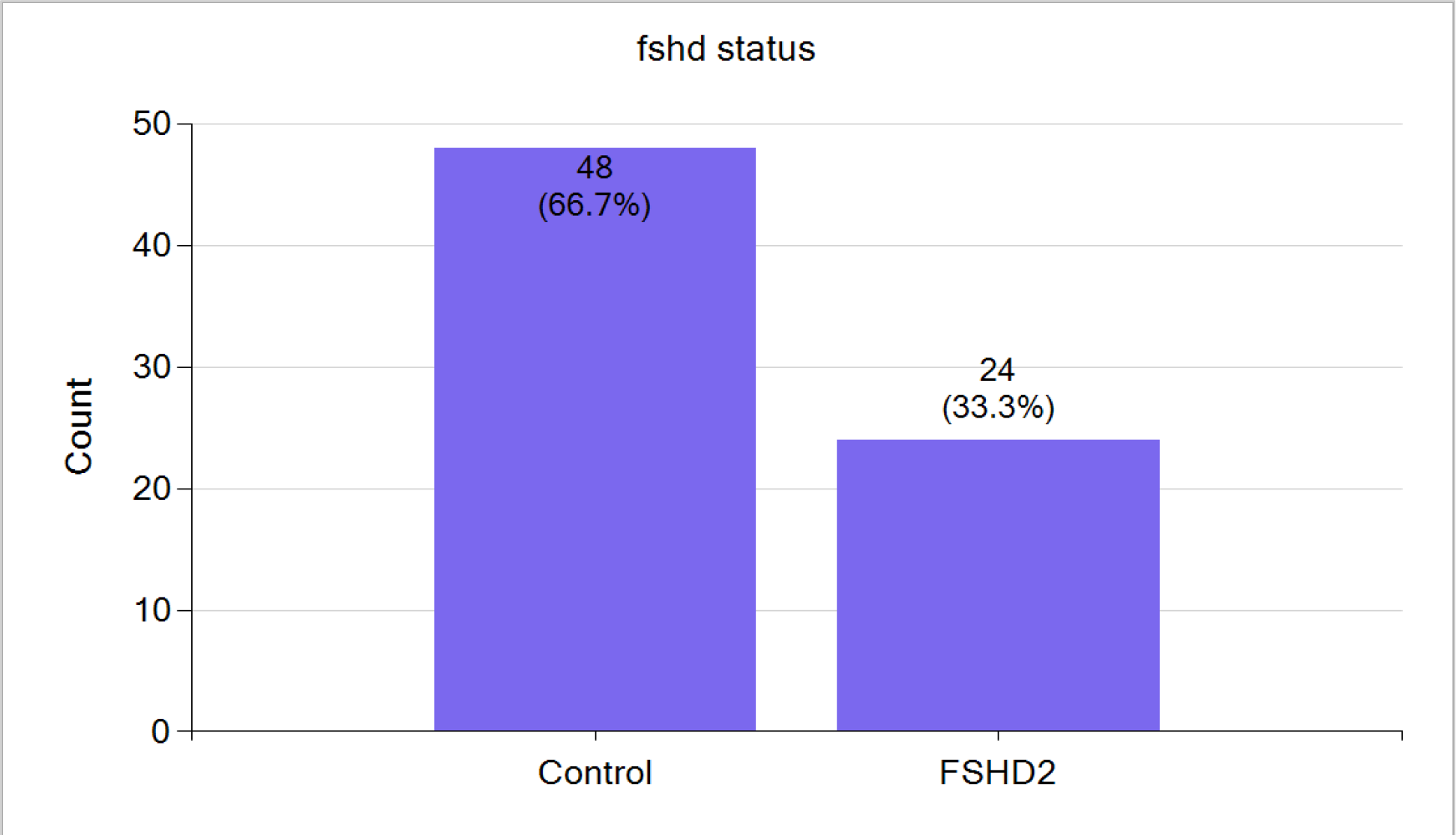
*GSE143453* comprises 48 control samples and 24 samples from individuals with FSHD2. The total number of genes is 10,827.

**Figure 2:**
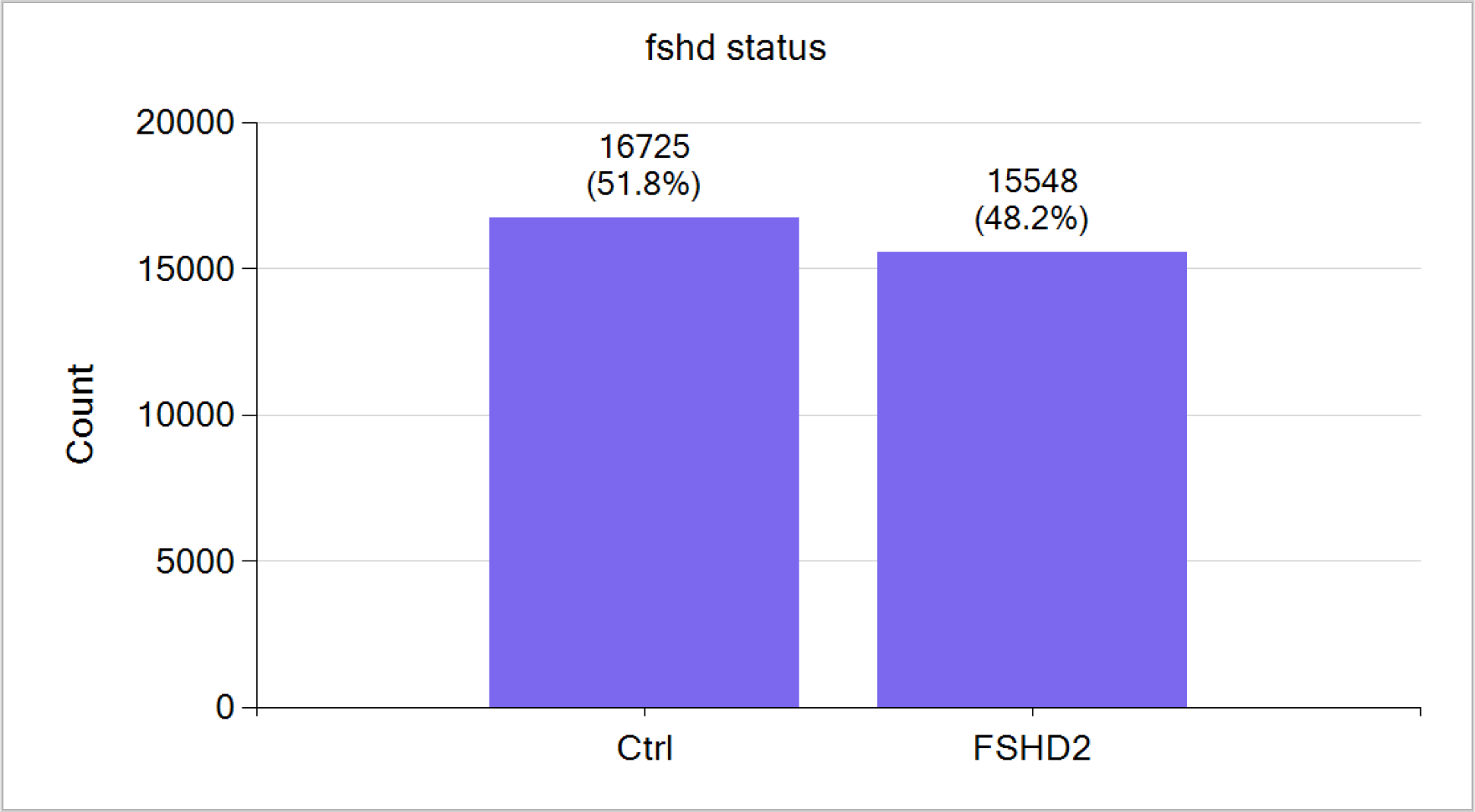
*GSE143452* comprises 16,725 control single cells and 15,548 FSHD2 single cells. The total number of genes is 19,615.

All gene-related data has been sourced from the *GeneCards* website and Google search. The pathway data from the *Reactome* website and the gene ontology data from the *QuickGO* website.

## Data Analysis

We conducted a comparative analysis between FSHD2 bulk and single-cell transcriptomes. Bulk transcriptomics provides an overall picture of gene expression in a sample, while single-cell transcriptomics allows for the identification of gene expression patterns at the individual cell level. These genes provide valuable insights into specific cell types and their unique gene expression profiles, making single-cell transcriptomics a valuable tool for dissecting cellular heterogeneity and understanding various biological processes. In this study, we analyze FSHD2 bulk and single-cell transcriptomes across three key categories: differential gene expressions, pathway ontology, and gene ontology.

### Differential Gene Expression – Up-regulated Genes

The top 10 up-regulated genes identified in the bulk gene expression analysis (**Table 1**) represent a diverse range of biological functions, but all are associated with FSHD. *MBD3L3* and *MBD3L2* are linked to DNA methylation regulation [2]. *RFPL4B* is predicted to enable metal ion binding activity. *TRIM43* is a member of the tripartite motif family known for its roles in immune response and protein degradation. *KHDC1L* is predicted to be involved in the apoptotic process. *LEUTX* is predicted to enable DNA-binding transcription factor activity. The protein encoded by *SLC34A2* is a pH-sensitive sodium-dependent phosphate transporter [3]. *ZSCAN4* contains domains relevant to gene expression regulation, while *TRIM43B* is a related gene with likely overlapping functions with *TRIM43* in immune and cellular regulation. Finally, *PRAMEF8* is predicted to be involved in several processes, including negative regulation of apoptotic process, negative regulation of DNA-templated transcription, and positive regulation of cell-population proliferation [4].

**Table 1:**
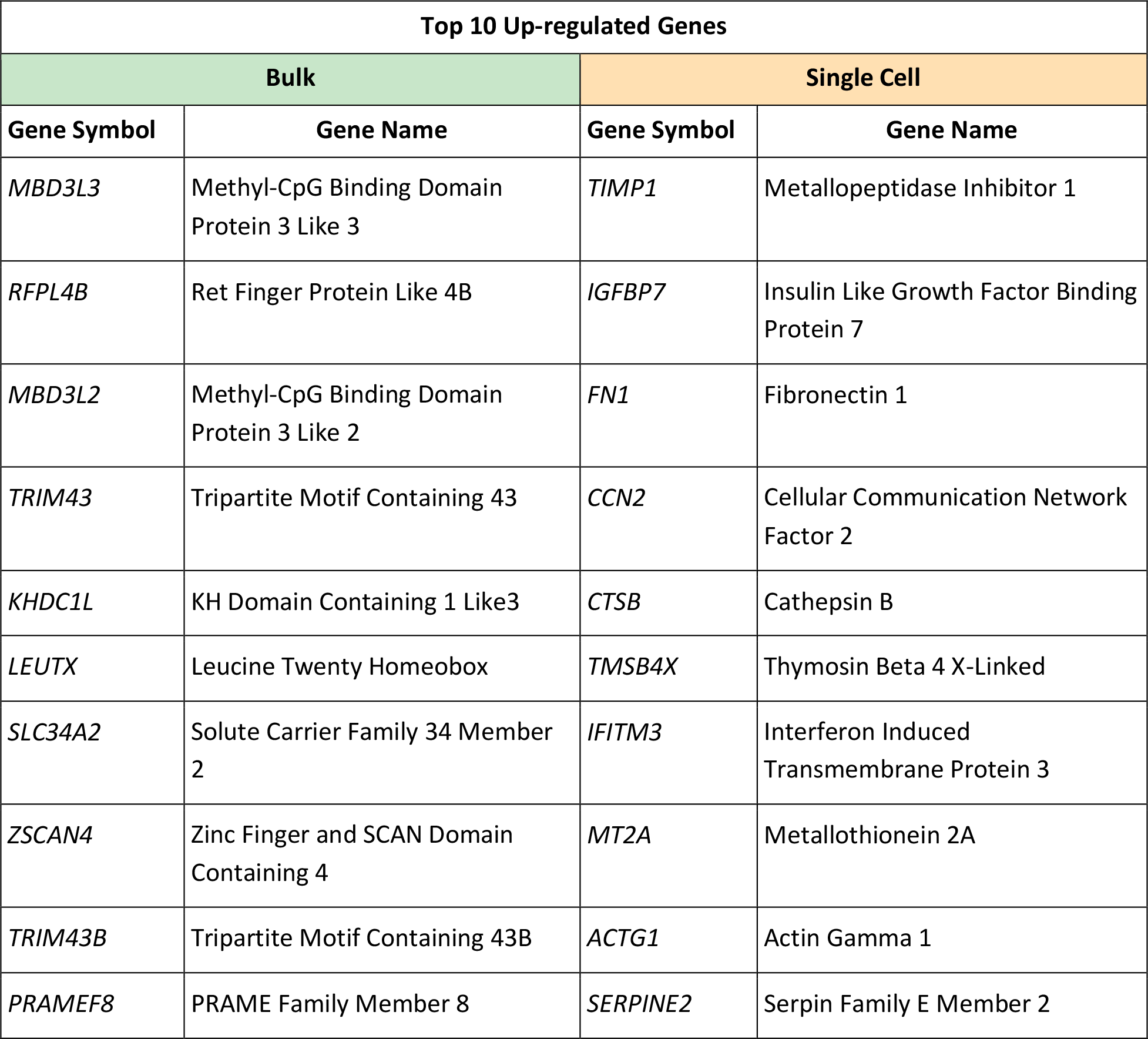
Top 10 up-regulated genes through bulk and single-cell gene expression analysis.

| Top 10 Up-regulated Genes |  |  |  |
| --- | --- | --- | --- |
| Bulk |  | Single Cell |  |
| Gene Symbol | Gene Name | Gene Symbol | Gene Name |
| *MBD3L3* | Methyl-CpG Binding Domain Protein 3 Like 3 | *TIMP1* | Metalloproteinase Inhibitor 1 |
| *RFPL4B* | Ret Finger Protein Like 4B | *IGFBP7* | Insulin Like Growth Factor Binding Protein 7 |
| *MBD3L2* | Methyl-CpG Binding Domain Protein 3 Like 2 | *FN1* | Fibronectin 1 |
| *TRIM43* | Tripartite Motif Containing 43 | *CCN2* | Cellular Communication Network Factor 2 |
| *KHDC1L* | KH Domain Containing 1 Like3 | *CTSB* | Cathepsin B |
| *LEUTX* | Leucine Twenty Homeobox | *TMSB4X* | Thymosin Beta 4 X-Linked |
| *SLC34A2* | Solute Carrier Family 34 Member 2 | *IFITM3* | Interferon Induced Transmembrane Protein 3 |
| *ZSCAN4* | Zinc Finger and SCAN Domain Containing 4 | *MT2A* | Metallothionein 2A |
| *TRIM43B* | Tripartite Motif Containing 43B | *ACTG1* | Actin Gamma 1 |
| *PRAMEF8* | PRAME Family Member 8 | *SERPINE2* | Serpin Family E Member 2 |

The top 10 upregulated genes identified in the single-cell gene expression analysis (**Table 1**) includes *TIMP1* which is involved in regulating metalloproteinase activity and the extracellular matrix [5]. *IGFBP7* is linked to insulin-like growth factor pathways and cell growth regulation [6]. *FN1* is involved in cell-adhesion and migration processes including embryogenesis, wound healing, blood coagulation, host defense, and metastasis [7]. *CCN2* is a mitogen secreted by vascular endothelial cells and plays a role in chondrocyte proliferation and differentiation, as well as cell adhesion in many cell types [8]. *CTSB* may initiate proteolytic pathways crucial for inflammatory breast cancer invasion and a potential prognostic marker for lymphatic metastasis [9]. *TMSB4X* encodes an actin-sequestering protein that plays a role in the regulation of actin polymerization. It is also involved in cell proliferation, migration, and differentiation. *IFITM3* regulates fibrinogen endocytosis and platelet reactivity in nonviral sepsis. The function of *MT2A* is to regulate metal homeostasis, detoxification, oxidative stress, immune defense, cell cycle progression, cell proliferation and differentiation, and angiogenesis [10]. *ACTG1* is a protein crucial for muscle contraction. Alterations in actin expression could directly impact muscle function and integrity [11]. *SERPINE2* encodes a member of the serpin family of proteins, a group of proteins that inhibit serine proteases [12].

As evident from the data presented in **Table 1**, any common genes within the top 10 up-regulated genes are distinctly absent when comparing bulk and single-cell gene expression. This disparity is anticipated given that bulk transcriptomics captures the collective gene expression within a cell population, while single-cell transcriptomics provides gene expression profiles for individual cells. Consequently, because of substantial disparities between these two distinct methodologies, the decision regarding which method to employ should be based on the specific research objectives and desired level of granularity.

### Differential Gene Expression – Down-regulated Genes

Several of the bulk top 10 down-regulated genes (**Table 2**) are related to muscular and cellular functions in some way. Notable genes include *SERF1B*, a positive regulator of amyloid protein aggregation and proteotoxicity and associated with Spinal Muscular Atrophy (SMA) Type III. *FLVCR1-DT* encodes a protein involved in heme transport and may be associated with anemias and other diseases. *SMN2* is a centromeric paralog of the *SMN1* gene. Mutations in *SMN1* are associated with SMA. *SMN2* produces a less functional form of the SMN protein, contributing to the disease’s severity [13]. *RTCA* encodes an enzyme involved in RNA metabolism and may play a role in RNA processing and stability. *SNHG5* is a long non-coding RNA that may regulate gene expression by acting as a sponge for microRNAs. *ACTR10* is associated with actin, a protein that plays a fundamental role in cell structure and movement. *DSTN* is a gene that encodes a protein involved in actin depolymerization, which is important for dynamic changes in the actin cytoskeleton. *CDC16* is one of several subunits of the anaphase-promoting complex, which functions at the metaphase-to-anaphase transition of the cell cycle and is regulated by spindle checkpoint proteins. *BBS2* is a gene associated with Bardet-Biedl syndrome, a rare genetic disorder that affects various body systems through its related pathways of organelle biogenesis and maintenance and cargo trafficking to the periciliary membrane. Finally, *SKAP2* is associated with signal transduction and may be involved in cell signaling pathways [14].

**Table 2:** Top 10 down-regulated genes through bulk and single-cell gene expression analysis.

| Top 10 Down-regulated Genes |  |  |  |
| --- | --- | --- | --- |
| Bulk |  | Single Cell |  |
| Gene Symbol | Gene Name | Gene Symbol | Gene Name |
| *SERF1B* | Small EDRK-Rich Factor 1B | *ACTC1* | Actin Alpha Cardiac Muscle 1 |
| *FLVCR1-DT* | FLVCR1 Divergent Transcript | *ACTA1* | Actin Alpha 1, Skeletal Muscle |
| *SMN2* | Survival Of Motor Neuron 2, Centromeric | *MYL11* | Myosin Light Chain 11 |
| *RTCA* | RNA 3'-Terminal Phosphate Cyclase | *MYH3* | Myosin Heavy Chain 3 |
| *SNHG5* | Small Nucleolar RNA Host Gene 5 | *MYL6B* | Myosin Light Chain 6B |
| *ACTR10* | Actin Related Protein 10 | *MYBPH* | Myosin Binding Protein H |
| *DSTN* | Destrin, Actin Depolymerizing Factor | *TPM2* | Tropomyosin 2 |
| *CDC16* | Cell Division Cycle 16 | *MYL2* | Myosin Light Chain 2 |
| *BBS2* | Bardet-Biedl Syndrome 2 | *MYL1* | Myosin Light Chain 1 |
| *SKAP2* | Src Kinase Associated Phosphoprotein 2 | *TNNI1* | Troponin I1, Slow Skeletal Type |

In contrast to the bulk down-regulated genes, the single-cell top 10 down-regulated genes (**Table 2**) are primarily linked to muscle-related functions. These genes—such as *ACTC1, ACTA1, MYL11, MYH3, MYL6B, MYBPH, TPM2, MYL2, MYL1*, and *TNNI1*—are integral to muscle contraction and skeletal muscle function, underscoring the precision and cell-specific nature of gene regulation in muscle tissues [15]. Even though none of these top 10 genes can be linked directly to FSHD, single-cell transcriptomics clearly represents a superior approach for discerning the top genes that distinguish the control group from the FSHD2 group.

However, further research is necessary to discern the exact implications of these gene expression changes in the context of specific biological functions and cellular responses.

### Pathway Analysis – Up-regulated Pathways

Several of the bulk top 10 down-regulated pathways (**Table 3**) can be linked to muscular dystrophy. In particular, the upregulation of *Type II Na+/Pi cotransporters* and *sodium-coupled phosphate cotransporters* may lead to imbalances in phosphate homeostasis, which can contribute to muscle degeneration. Additionally, abnormalities in *surfactant metabolism*, such as those seen in diseases associated with surfactant metabolism and defective SLC34A2 causing pulmonary alveolar microlithiasis (PALM), can disrupt lung function, leading to respiratory complications common in muscular dystrophy. *G2 Phase* and *phosphorylation of proteins involved in the G2/M transition by Cyclin A:Cdc2 complexes* may impact cell cycle regulation, potentially influencing muscle cell growth and repair processes. Furthermore, *regulation of TP53*, including its expression, transcriptional activity, and degradation, plays a critical role in controlling cell cycle progression and apoptosis, which are essential processes in muscle maintenance and repair. Thus, these upregulated pathways, when dysregulated, can contribute to the pathophysiology of muscular dystrophy [16].

**Table 3:** Top 10 up-regulated pathways through bulk and single-cell gene expression analysis.

| Top 10 Up-regulated Pathways |  |
| --- | --- |
| Bulk | Single Cell |
| Type II Na<sup>+</sup>/Pi cotransporters | Interleukin-10 signaling |
| Defective SLC34A2 causes pulmonary alveolar microlithiasis (PALM) | Activation of Matrix Metalloproteinases |
| Diseases associated with surfactant metabolism | Post-translational protein phosphorylation |
| Sodium-coupled phosphate cotransporters | Regulation of Insulin-like Growth Factor (IGF) transport and uptake by Insulin-like Growth Factor Bi |
| G2 Phase | Cytokine Signaling in Immune system |
| Phosphorylation of proteins involved in the G2/M transition by Cyclin A:Cdc2 complexes | Signaling by Interleukins |
| TP53 Regulates Transcription of Genes Involved in G1 Cell Cycle Arrest | Interleukin-4 and Interleukin-13 signaling |
| Regulation of TP53 Expression and Degradation | MAP2K and MAPK activation |
| Regulation of TP53 Degradation | Signaling by high-kinase activity BRAF mutants |
| surfactant metabolism | Signaling by moderate kinase activity BRAF mutants |

Among the top 10 up-regulated pathways for the single-cell transcriptome (**Table 3**) there are many pathways related to muscular dystrophy. First, the upregulation of *Activation of Matrix Metalloproteinases* may lead to increased degradation of extracellular matrix components in muscle tissue, contributing to muscle damage. Additionally, *Interleukin-10 signaling* and *Cytokine Signaling in the Immune system* suggest the involvement of inflammation, which can exacerbate muscle degeneration. *MAP2K and MAPK activation* pathways may play a role in regulating cell growth and differentiation, potentially influencing muscle tissue maintenance. *Regulation of Insulin-like Growth Factor (IGF) transport and uptake by Insulin-like Growth Factor Binding Proteins* can affect the availability of growth factors critical for muscle repair and maintenance. Overall, the dysregulation of these pathways could contribute to the pathophysiology of FSHD2 by promoting inflammation, impairing muscle regeneration, and disrupting muscle tissue homeostasis.

### Pathway Analysis – Down-regulated Pathways

Several of the top 10 bulk down-regulated pathways (**Table 4**) are implicated in the pathogenesis of muscle dystrophy. Downregulation of *Calcineurin activates NFAT* reduces the activation of nuclear factor of activated T-cells (NFAT), which plays a role in muscle development and regeneration. *FCERI mediated Ca+2 mobilization* pertains to calcium mobilization, an essential process for muscle contraction and relaxation. *Synthesis of IP3 and IP4 in the cytosol* is involved in intracellular calcium regulation, crucial for muscle function. *CLEC7A (Dectin-1) induces NFAT activation* is related to immune responses that can impact muscle health. *Antigen Presentation: Folding, assembly and peptide loading of class I MHC* is essential for immune surveillance in muscle tissue. *Dectin-2 family* and *Arachidonate production from DAG* can influence immune responses and inflammation in muscles. *Sodium/Calcium exchangers* are integral to calcium homeostasis in muscle cells. *COPI-mediated anterograde transport* and *ER to Golgi Anterograde Transport* relate to intracellular trafficking, which is crucial for proper protein synthesis and muscle maintenance.

**Table 4:** Top 10 down-regulated pathways through bulk and single-cell gene expression analysis.

| Top 10 Down-regulated Pathways |  |
| --- | --- |
| Bulk | Single Cell |
| Calcineurin activates NFAT | Striated Muscle Contraction |
| FCERI mediated Ca<sup>2+</sup> mobilization | Muscle contraction |
| Synthesis of IP3 and IP4 in the cytosol | Smooth Muscle Contraction |
| CLEC7A (Dectin-1) induces NFAT activation | Creatine metabolism |
| Antigen Presentation: Folding, assembly and peptide loading of class I MHC | Assembly and cell surface presentation of NMDA receptors |
| Dectin-2 family | Kinesins |
| Sodium/Calcium exchangers | Carboxyterminal post-translational modifications of tubulin |
| COPI-mediated anterograde transport | Post-chaperonin tubulin folding pathway |
| Arachidonate production from DAG | MECP2 regulates transcription factors |
| ER to Golgi Anterograde Transport | Myogenesis |

All the top 10 single-cell down-regulated pathways (**Table 4**) are implicated in the pathogenesis of muscle dystrophy. These pathways include down-regulation of *striated muscle contraction, muscle contraction* (both essential for proper muscle function), and *myogenesis* (critical for muscle development and regeneration). Additionally, the reduced activity of *smooth muscle contraction* may affect overall muscle integrity. Furthermore, disruptions in *creatine metabolism* can lead to energy deficits in muscle cells. The down-regulation of *assembly and cell surface presentation of NMDA receptors* may disrupt neuromuscular signaling, while the decreased activity of *kinesins, carboxyterminal post-translational modifications of tubulin*, and *post-chaperonin tubulin folding pathway* can impair the structural integrity and intracellular transport of muscle proteins. The altered *regulation of transcription factors by MECP2* may further contribute to muscle dysfunction in FSHD2. Overall, these downregulated pathways collectively play a role in the complex etiology of FSHD2, impacting muscle function, structure, and development.

The bulk downregulated pathways provide insights into a broader range of potential mechanisms involved in FSHD2, including immune-related pathways, calcium regulation, and intracellular trafficking. These insights can be valuable in understanding the disease’s complexity and its systemic impact. On the other hand, the single-cell down-regulated pathways delve deeper into the specific pathways critical for muscle function, development, and integrity, offering a more detailed view of how these factors contribute to muscle dysfunction in FSHD2.

### Gene Ontology Analysis – Up-regulated Biological Process (BP), Cellular Component (CC) or Molecular Function (MF)

Several gene ontology-related items (bulk) can be linked to muscular dystrophy in general (**Table 5**). The up-regulation of genes associated with *methylation-dependent chromatin silencing* could contribute to altered gene expression patterns in muscle cells, potentially exacerbating the disease. *Negative regulation of mitotic recombination* may impede the repair of damaged DNA in muscle cells, which could be involved in the pathogenesis of muscular dystrophy. Additionally, *cellular phosphate ion homeostasis* and *sodium-dependent phosphate transport* are important for muscle function, and their upregulation may indicate a compensatory response in affected muscles. *Negative regulation of cell differentiation* could hinder muscle regeneration, while *positive regulation of cysteine-type endopeptidase activity involved in execution phase of apoptosis* may lead to muscle cell death. Lastly, *telomere maintenance via telomere lengthening* may be implicated in muscle cell aging and degeneration in muscular dystrophy. The *cyclin A1-CDK2 complex* and *sodium:phosphate symporter activity* might indirectly influence these processes through their roles in cell cycle regulation and phosphate transport.

**Table 5:**
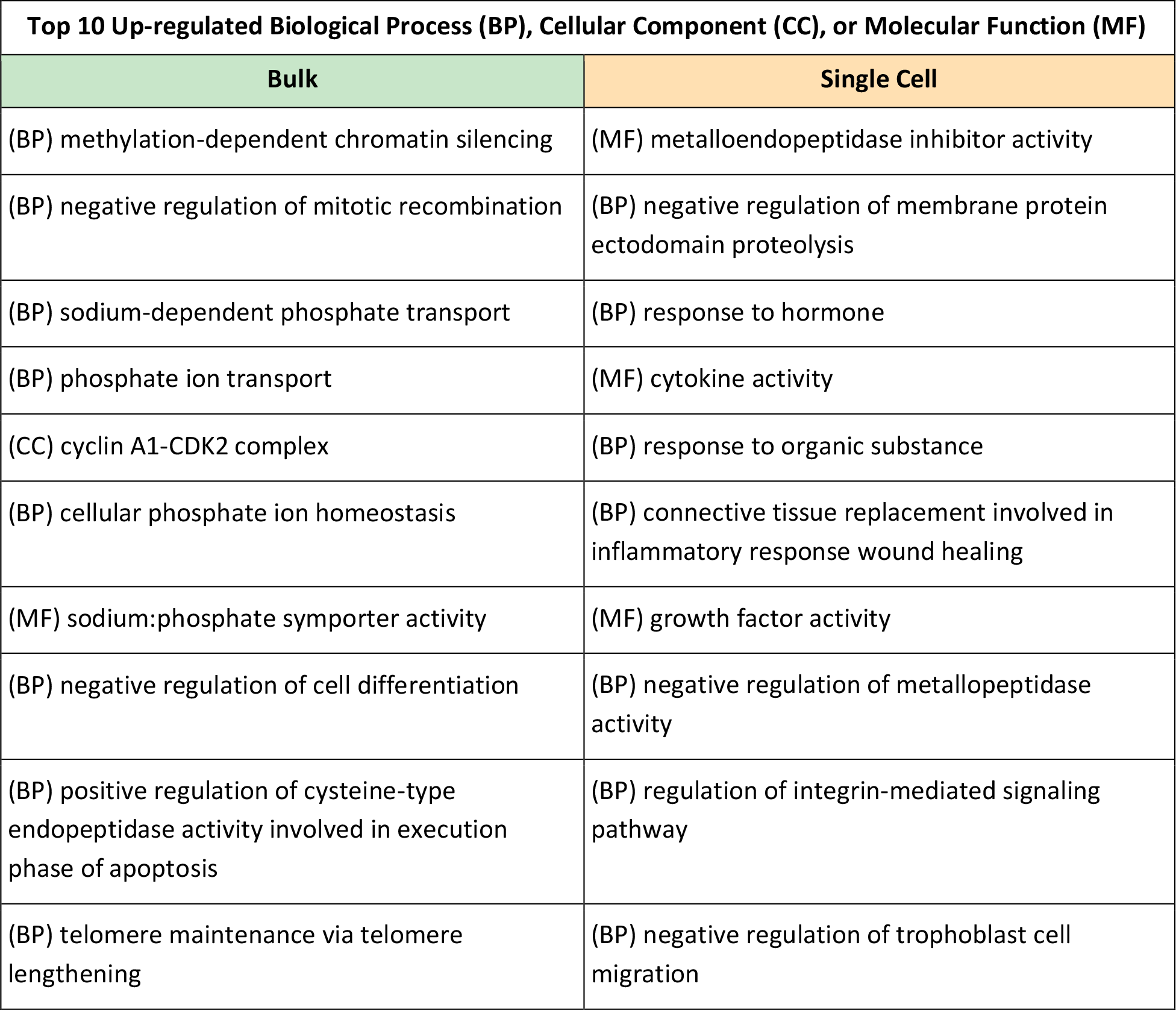
Top 10 upregulated gene ontology terms through bulk and single-cell gene expression analysis.

Several up-regulated gene ontology-related processes and molecular functions (single-cell) are implicated (**Table 5**). First, *negative regulation of membrane protein ectodomain proteolysis* indicates an alteration in the regulation of protease activity, which could influence muscle membrane integrity. Additionally, *negative regulation of metallopeptidase activity* and metallopeptidase inhibitor activity both suggest an imbalance in metalloendopeptidase activity, which may impact muscle tissue. *Response to hormone* signifies potential hormonal disruptions affecting muscle homeostasis. *Cytokine activity* and *growth factor activity* imply an altered inflammatory and growth response in the context of muscle repair. Furthermore, *connective tissue replacement involved in inflammatory response wound healing* points to the involvement of inflammatory processes in muscle tissue repair and fibrosis. *Regulation of integrin-mediated signaling pathway* suggests alterations in cell adhesion and signaling, which can impact muscle function. Finally, *negative regulation of trophoblast cell migration* hints at disrupted cell migration, possibly affecting muscle regeneration.

### Gene Ontology Analysis – Down-regulated BP, CC, or MF

Several of the down-regulated gene ontology terms (**Table 6**) may have relevance to muscular dystrophy. Down-regulation of *NADPH:quinone reductase activity* can impair the antioxidant defense system, leading to increased oxidative stress and muscle damage. Reduced *retinol dehydrogenase activity* may impact retinoid metabolism, which is important for muscle regeneration and repair. Additionally, the downregulation of *protein-disulfide reductase activity* may contribute to protein misfolding and aggregation, a hallmark of some forms of muscular dystrophy. These molecular changes could disrupt various biological processes including *negative regulation of hepatocyte proliferation* which might exacerbate muscle damage, and *antigen processing and presentation of peptide antigen via MHC class I*, which could influence immune responses in muscle tissues. Furthermore, *regulation of synaptic vesicle endocytosis* may be altered, impacting neuromuscular communication, and *retrograde axonal transport of mitochondrion* may lead to mitochondrial dysfunction, which is associated with muscle weakness.

**Table 6:** Top 10 down-regulated gene ontology terms through bulk and single-cell gene expression analysis.

| Top 10 Down-regulated BP, CC, or MF |  |
| --- | --- |
| Bulk | Single Cell |
| (MF) NADPH:quinone reductase activity | (BP) cardiac muscle contraction |
| (MF) RNA-3-phosphate cyclase activity | (BP) muscle filament sliding |
| (BP) positive regulation of epithelial cell proliferation involved in lung morphogenesis | (BP) skeletal muscle thin filament assembly |
| (BP) retrograde axonal transport of mitochondrion | (BP) mesenchyme migration |
| (MF) retinol dehydrogenase activity | (CC) sarcomere |
| (BP) negative regulation of hepatocyte proliferation | (MF) myosin binding |
| (BP) antigen processing and presentation of peptide antigen via MHC class I | (CC) Isotropic band (I band) |
| (BP) regulation of synaptic vesicle endocytosis | (BP) muscle contraction |
| (BP) viral budding | (BP) cardiac myofibril assembly |
| (MF) protein-disulfide reductase activity | (BP) cardiac muscle tissue morphogenesis |

The top 10 down-regulated gene ontology terms are all relevant to the pathogenesis of muscular dystrophy. In individuals with this condition, *cardiac muscle contraction* and *muscle contraction* in general are significantly down-regulated, resulting in weakened muscular function. This enfeeblement is accompanied by a decrease in *muscle filament sliding* and *skeletal muscle thin filament assembly*, essential processes for muscle strength. Additionally, the down-regulation of *sarcomere, Isotropic band (I band)*, and *myosin binding* suggests structural and functional abnormalities in the muscle fibers, contributing to muscle weakness. Furthermore, reduced *cardiac myofibril assembly* and *cardiac muscle tissue morphogenesis* indicate impaired development and maintenance of cardiac muscle, which is often affected in muscular dystrophy. Lastly, the down-regulation of *mesenchyme migration* may reflect disruptions in muscle cell migration and differentiation. Altogether, these down-regulated gene ontology terms underscore the molecular mechanisms involved in the development and progression of FSHD2.

## Discussion

This paper focuses on exploring the molecular aspects of FSHD, a genetic muscle disorder characterized by progressive muscle weakening, particularly in the face, shoulder blades, and upper arms. The two types of FSHD, types 1 and 2, have distinct genetic causes. FSHD1 is associated with a chromosome-4 deletion, while FSHD2 results from mutations in the *SMCHD1* gene, leading to the activation of the *DUX4* gene in muscle cells, causing premature cell death. The study employs bulk and single-cell transcriptomics to investigate the gene expression patterns associated with FSHD2. Bulk transcriptomics provides an overall view of gene expression in a sample, while single-cell transcriptomics offers insights into individual cell-level gene expression. The analysis covers three main categories: differential gene expression, pathway analysis, and gene ontology analysis, primarily in the context of FSHD2.

The top 10 up-regulated genes in bulk transcriptomics are associated with various biological functions, all relevant to FSHD. In contrast, the top 10 up-regulated genes in single-cell transcriptomics are primarily linked to muscle-related functions, indicating a cell-specific nature of gene regulation. In bulk transcriptomics, down-regulated genes have diverse functions, including immune-related processes, calcium regulation, and intracellular trafficking. In single-cell transcriptomics, down-regulated genes are predominantly related to muscle function, emphasizing their role in muscle dysfunction in FSHD2.

Upregulated pathways in bulk transcriptomics relate to muscle dystrophy and include processes involving phosphate homeostasis, surfactant metabolism, and cell cycle regulation. Single-cell transcriptomics reveal pathways associated with inflammation, extracellular matrix degradation, and growth factor regulation, impacting muscle tissue homeostasis. Bulk transcriptomics down-regulated pathways encompass immune responses, calcium regulation, and intracellular trafficking. In contrast, single-cell transcriptomics down-regulated pathways are more specific to muscle function, development, and structural integrity.

Up-regulated gene ontology terms in bulk transcriptomics indicate potential disruptions in gene-expression patterns, cell differentiation, phosphate transport, and apoptosis. In single-cell transcriptomics, up-regulated terms suggest alterations in protease activity, inflammatory and growth responses, cell adhesion, and cell migration, all related to muscle tissue. Down-regulated terms in bulk transcriptomics may affect antioxidant defense, retinoid metabolism, protein folding, and immune responses in muscle. In single-cell transcriptomics, down-regulated terms highlight the significant impact on muscle contraction, muscle filament assembly, muscle structure, and cardiac muscle development.

Overall, this study provides a comprehensive analysis of gene-expression patterns, pathways, and gene ontology terms associated with FSHD2, emphasizing the complex molecular mechanisms involved in the disease’s development and progression.

### Summary

This study on FSHD utilizes both bulk and single-cell transcriptomics to investigate gene-expression patterns in FSHD2. The key superiority of single-cell transcriptomics becomes evident in its ability to reveal cell-specific gene regulation, particularly in muscle-related functions, and to identify down-regulated genes primarily related to muscle dysfunction, while bulk transcriptomics provides a more general view of gene expression. Single-cell transcriptomics also highlights pathways and gene ontology terms specific to muscle tissue, emphasizing the complex molecular mechanisms involved in FSHD2, making it a valuable tool in understanding the disease at a finer level of detail.

## Data Availability

All data produced are available online at:
https://www.ncbi.nlm.nih.gov/geo/query/acc.cgi?acc=GSE143452
https://www.ncbi.nlm.nih.gov/geo/query/acc.cgi?acc=GSE143453

https://www.ncbi.nlm.nih.gov/geo/query/acc.cgi?acc=GSE143452

## References

1. Schätzl, T., Kaiser, L. & Deigner, HP. Facioscapulohumeral muscular dystrophy: genetics, gene activation and downstream signalling with regard to recent therapeutic approaches: an update. Orphanet J Rare Dis 16, 129 (2021).

2. Jin SG, Jiang CL, Rauch T, Li H, Pfeifer GP. MBD3L2 interacts with MBD3 and components of the NuRD complex and can oppose MBD2-MeCP1-mediated methylation silencing. J Biol Chem. 2005 Apr 1;280(13):12700–9.

3. Mueller AL, O’Neill A, Jones TI, Llach A, Rojas LA, Sakellariou P, Stadler G, Wright WE, Eyerman D, Jones PL, Bloch RJ. Muscle xenografts reproduce key molecular features of facioscapulohumeral muscular dystrophy. Exp Neurol. 2019 Oct;320:113011.

4. Lim KRQ, Bittel A, Maruyama R, Echigoya Y, Nguyen Q, Huang Y, Dzierlega K, Zhang A, Chen YW, Yokota T. DUX4 Transcript Knockdown with Antisense 2’-O-Methoxyethyl Gapmers for the Treatment of Facioscapulohumeral Muscular Dystrophy. Mol Ther. 2021 Feb 3;29(2):848–858.

5. Koutsoulidou A, Phylactou LA. Circulating Biomarkers in Muscular Dystrophies: Disease and Therapy Monitoring. Mol Ther Methods Clin Dev. 2020 May 22;18:230–239.

6. Alsulaiman SM, Abu-Safieh L, AlJarallah AS, AlAbdulhafid M, AlKahtani ES. Advanced coats-like retinopathy as the initial presentation of Familial Retinal Arterial Macroaneurysms. Am J Ophthalmol Case Rep. 2018 Apr 17;11:153–157.

7. Laberthonniére C, Novoa-Del-Toro EM, Delourme M, Chevalier R, Broucqsault N, Mazaleyrat K, Streichenberger N, Manel V, Bernard R, Salort Campana E, Attarian S, Nguyen K, Robin JD, Baudot A, Magdinier F. Facioscapulohumeral dystrophy weakened sarcomeric contractility is mimicked in induced pluripotent stem cells-derived innervated muscle fibres. J Cachexia Sarcopenia Muscle. 2022 Feb;13(1):621–635.

8. Petrosino JM, Leask A, Accornero F. Genetic manipulation of CCN2/CTGF unveils cell-specific ECM-remodeling effects in injured skeletal muscle. FASEB J. 2019 Feb;33(2):2047–2057.

9. Celegato B, Capitanio D, Pescatori M, Romualdi C, Pacchioni B, Cagnin S, Viganó A, Colantoni L, Begum S, Ricci E, Wait R, Lanfranchi G, Gelfi C. Parallel protein and transcript profiles of FSHD patient muscles correlate to the D4Z4 arrangement and reveal a common impairment of slow to fast fibre differentiation and a general deregulation of MyoD-dependent genes. Proteomics. 2006 Oct;6(19):5303–21.

10. Sara T. Winokur, Yi-Wen Chen, Peter S. Masny, Jorge H. Martin, Jeffrey T. Ehmsen, Stephen J. Tapscott, Silvere M. van der Maarel, Yukiko Hayashi, Kevin M. Flanigan, Expression profiling of FSHD muscle supports a defect in specific stages of myogenic differentiation, Human Molecular Genetics, Volume 12, Issue 22, 15 November 2003, Pages 2895–2907.

11. Liu Q, Jones TI, Tang VW, Brieher WM, Jones PL. Facioscapulohumeral muscular dystrophy region gene-1 (FRG-1) is an actin-bundling protein associated with muscle-attachment sites. J Cell Sci. 2010 Apr 1;123(Pt 7):1116–23.

12. Tjondrokoesoemo A, Schips T, Kanisicak O, Sargent MA, Molkentin JD. Genetic overexpression of Serpina3n attenuates muscular dystrophy in mice. Hum Mol Genet. 2016 Mar 15;25(6):1192–202. doi: 10.1093/hmg/ddw005. Epub 2016 Jan 6. PMID: 26744329; PMCID: PMC4764199.

13. Chiu W, Hsun YH, Chang KJ, Yarmishyn AA, Hsiao YJ, Chien Y, Chien CS, Ma C, Yang YP, Tsai PH, Chiou SH, Lin TY, Cheng HM. Current Genetic Survey and Potential Gene-Targeting Therapeutics for Neuromuscular Diseases. Int J Mol Sci. 2020 Dec 16;21(24):9589.

14. Tumasian RA 3rd, Harish A, Kundu G, Yang JH, Ubaida-Mohien C, Gonzalez-Freire M, Kaileh M, Zukley LM, Chia CW, Lyashkov A, Wood WH 3rd, Piao Y, Coletta C, Ding J, Gorospe M, Sen R, De S, Ferrucci L. Skeletal muscle transcriptome in healthy aging. Nat Commun. 2021 Apr 1;12(1):2014.

15. Rahimov F, King OD, Leung DG, Bibat GM, Emerson CP Jr, Kunkel LM, Wagner KR. Transcriptional profiling in facioscapulohumeral muscular dystrophy to identify candidate biomarkers. Proc Natl Acad Sci U S A. 2012 Oct 2;109(40):16234–9.

16. Rahimov F, King OD, Leung DG, Bibat GM, Emerson CP Jr, Kunkel LM, Wagner KR. Transcriptional profiling in facioscapulohumeral muscular dystrophy to identify candidate biomarkers. Proc Natl Acad Sci U S A. 2012 Oct 2;109(40):16234–9.

